# Immunohistochemistry and Next Generation Sequencing are Complementary Tests in Identifying PTEN Abnormality in Endometrial Carcinoma Biopsies

**DOI:** 10.1101/2020.07.20.20154641

**Authors:** Linyuan Wang, Anna Piskorz, Tjalling Bosse, Mercedes Jimenez-Linan, Brian Rous, C. Blake Gilks, James D. Brenton, Naveena Singh, Martin Köbel

**Affiliations:** Department of Pathology, University of Calgary, Calgary, Alberta, Canada; Cancer Research UK Cambridge Institute, University of Cambridge, UK; Department of Pathology, University of Leiden, Leiden, The Netherlands; Department of Pathology, Addenbrooke’s Hospital, Cambridge, UK; Department of Pathology Vancouver General Hospital, Vancouver, British Columbia, Canada; Department of Pathology, Barts Health NHS Trust, London, UK

**Keywords:** Endometrial carcinoma, molecular subtype, PTEN

## Abstract

PTEN plays a central role in the pathogenesis of endometrial carcinoma. Previous studies reported a high interobserver reproducibility for the interpretation of PTEN immunohistochemistry (IHC). However, PTEN IHC and its interpretation remain challenging during laboratory practice. The purpose of this study was to reevaluate PTEN IHC pattern in direct comparison to next generation sequencing (NGS) in identifying PTEN abnormality. IHC and tagged-amplicon NGS *PTEN* sequencing was performed on 182 endometrial carcinoma biopsy/curetting samples from five centers (Barts, Calgary, Cambridge, Leiden, and Vancouver). Sensitivity, specificity and accuracy of PTEN IHC to predict loss of function (LOF) *PTEN* mutations were calculated. Abnormalities of PTEN in association with histotype and molecular subtype were assessed. A total of five PTEN IHC patterns were recorded: absent, subclonal loss, equivocal, reduced (relative to internal control) and retained. The absence of PTEN IHC has a sensitivity of 75.4% (95% CI 62.7 – 85.5%), a specificity of 84.6% (95% CI 76.2 – 90.9%), and accuracy of 81.2% (95% CI 74.4 – 86.9%) in predicting LOF *PTEN* mutation. PTEN abnormality by complementary interpretation of both assays was present in 91.9% of endometrial endometrioid carcinoma, grade 1, and significantly higher in endometrial endometrioid carcinomas of all grades compared to endometrial serous carcinoma (80.0% versus 19.4%, p<0.0001). PTEN abnormalities are common across all molecular subtypes of endometrioid carcinomas. Our data support complementary testing of both IHC and sequencing of PTEN to assess the PTEN status in endometrial carcinomas.

## INTRODUCTION

*PTEN* is the most commonly altered gene in endometrial endometrioid carcinoma (84%) (1). It is often an early event detected in its precursor atypical endometrial hyperplasia (2). Women with *PTEN* germline mutations are at increased risk for endometrial neoplasms as part of the *PTEN* hamartoma tumor syndrome (PHTS)/Cowden syndrome (3). Although there is considerable variation in mutational frequency reported in published series, *PTEN* alterations are characteristic in the carcinogenesis of the endometrioid histotype and they are uncommon in endometrial serous carcinomas (2.6% in the TCGA dataset) (1). We previously proposed the use of PTEN immunohistochemistry (IHC) in distinguishing endometrioid from serous histotype (4-7). Yet despite reports showing good interobserver reproducibility and value as surrogate test for *PTEN* mutational status, PTEN IHC is not widely used in diagnostic practice, in part due to the ill-defined interpretation criteria regarding the staining (8,9).

The treatment principle for endometrial carcinoma remains early detection followed by surgical excision with or without radio- and/or chemotherapy (10). However, for patients with advanced disease or recurrent disease, or those patients in favor of fertility preservation, systemic therapy targeting the PI3K/AKT/mTOR pathway has showed promising result in various stages of clinical trials (11). Thus, reliable identification of PTEN abnormality is not only useful in the correct histotype diagnosis of endometrial carcinoma but relevant for clinical trial inclusion.

Two contrasting IHC patterns have been well-accepted and confirmed by multiple publications: the normal “Retained” pattern with PTEN expression in the cytoplasm of tumor cells; and the “Absent/Complete loss” pattern characterized by no PTEN staining in tumor cells but retained expression in stromal (often nuclear) and inflammatory cells which serves as internal control. In addition, two other staining patterns have been reported. The “Sub-clonal loss/Heterogeneous” pattern shows geographic loss of PTEN protein expression with areas of retained staining. Another “Reduced/Attenuated” pattern shows a lower intensity of PTEN expression in tumor cells compared to the internal controls. Both sub-clonal loss and reduced staining have been previously interpreted as abnormal PTEN expression (2,9,12), however, the correlation of the latter two staining patterns with mutational analysis has not been well characterized.

The aim of this study is to assess how accurate whole section PTEN IHC predicts *PTEN* loss of function (LOF) mutations assessed by next generation sequencing in a large cohort of endometrial biopsy specimens. A secondary aim was to evaluate association of PTEN alterations with histotype and molecular subtype in endometrial carcinomas.

## MATERIAL AND METHODS

### Patient samples

Sections from endometrial carcinoma biopsy/curettage samples collected for a previous study were used. Specimens were collected from five pathology departments and included cases with a diagnosis of endometrial carcinoma serous:non-serous high grade:G1-2 endometrioid EC in an approximately 2:1:1 ratio with the goal of identifying 200 cases with >50% prevalence of *TP53* mutation for the purpose of a previous study (13). PTEN IHC was carried out centrally using a DAKO Omnis protocol (H30-R10-30) and the rabbit monoclonal PTEN Cell Signaling clone 138G6 at a dilution of 1/60. Ethics approval was received from the Health Research Ethics Board of Alberta Cancer Committee (HREBA-CC-16-0156).

### PTEN IHC: Interpretation of Immunohistochemical Patterns

PTEN IHC was independently interpreted by two study pathologists (LW, MK) blinded to the mutation data. IHC discordant cases were revaluated at a multiheaded microscope for consensus. The following patterns recorded: 1) retained for similar PTEN IHC staining intensity in tumor cell cytoplasm compared to the internal control; 2) complete absence of PTEN staining in tumor cells with retained internal control (intervening stroma and inflammatory cells); 3) subclonal loss with a clearly recognizable zone of complete absence of PTEN staining in tumor cells with retained internal control in this zone and other areas with unequivocal expression (retained or reduced), 4) reduced for unequivocal PTEN staining in tumor cytoplasm but less staining intensity compared to the internal control; 5) and a new equivocal category for cases with just weak cytoplasmic blush of PTEN staining in tumor cells, which remained difficult to classify even after consensus assessment (12). Discordant cases between the observers were finalized by consensus assessment at a multiheaded microscope. No revisions of the IHC scores were made after being unblinded to the mutational data.

### PTEN sequencing

Targeted tagged amplicon deep sequencing (TAm-Seq) as described [19] on an Illumina MiSeq or HiSeq4000 using PE-150bp protocol was performed for a previous study (13). The entire coding region for PTEN was included in the targeted panel. Sequencing data and variant verification were performed using an in-house analysis pipeline and IGV software as described previously (14). *PTEN* mutations were called using publicly available databases (ClinVAr and COSMIC (15,16).

### Statistical Analysis

*POLE* mutation, mismatch repair and p53 status data were available from a previous study (13) Sensitivity, specificity and agreement or accuracy with 95% confidence intervals were calculated.

## RESULTS

### PTEN IHC Interobserver Agreement and Concordance with NGS

Initially, 207 endometrial biopsy/curettage specimens with a diagnosis of endometrial carcinoma were collected from five centres. 24 cases failed sequencing and 1 case failed PTEN IHC, hence, 182 cases with PTEN sequencing and IHC data were included in the analysis. With our PTEN IHC protocol, we identified a subset of cases with a new staining pattern of PTEN with weak cytoplasmic blush, we termed equivocal category (Figure 1). When the cohort was stratified into abnormal IHC staining (complete absence and subclonal loss) versus others, the interobserver reproducibility was moderate (kappa 0.71, agreement in 86%, 156/182 cases). The distribution of consolidated IHC categories is shown in Table 1. Deleterious *PTEN* mutations (LOF or NSM) were detected in 42% of cases. The majority of cases with LOF (56.7%) or nonsynonymous mutations (77.8%) also harbored a second or even a third *PTEN* mutations. The vast majority of the second or third *PTEN* mutations were nonsynonymous.

**Table 1:** Agreement between PTEN Immunohistochemistry and Next Generation Sequencing.

|  | LOF | NS | VUS | NMD | Total |
| --- | --- | --- | --- | --- | --- |
| Absent | 46 (74.2%) | 1 (1.6%) | 2 (3.2%) | 13 (21.0%) | 62 (34.1%) |
| Sub-clonal Loss | 6 (35.3%) | 0 (0%) | 2 (11.8%) | 9 (52.9%) | 17 (9.3%) |
| Equivocal | 4 (33.3%) | 0 | 0 | 8 (66.7%) | 12 (6.6%) |
| Reduced | 5 (12.8%) | 3 (7.7%) | 2 (5.1%) | 29 (74.4%) | 39 (21.4%) |
| Retained | 6 (11.5%) | 5 (9.6) | 6 (11.5%) | 35 (67.3%) | 52 (28.6%) |
| Total | 67 (36.8%) | 9 (5.0%) | 12 (6.6%) | 94 (51.6%) | 182 |

LOF:
loss of function, NS: non-synonymous, VUS: variance of uncertain significance, NMD: no mutation detected

**Figure 1.**
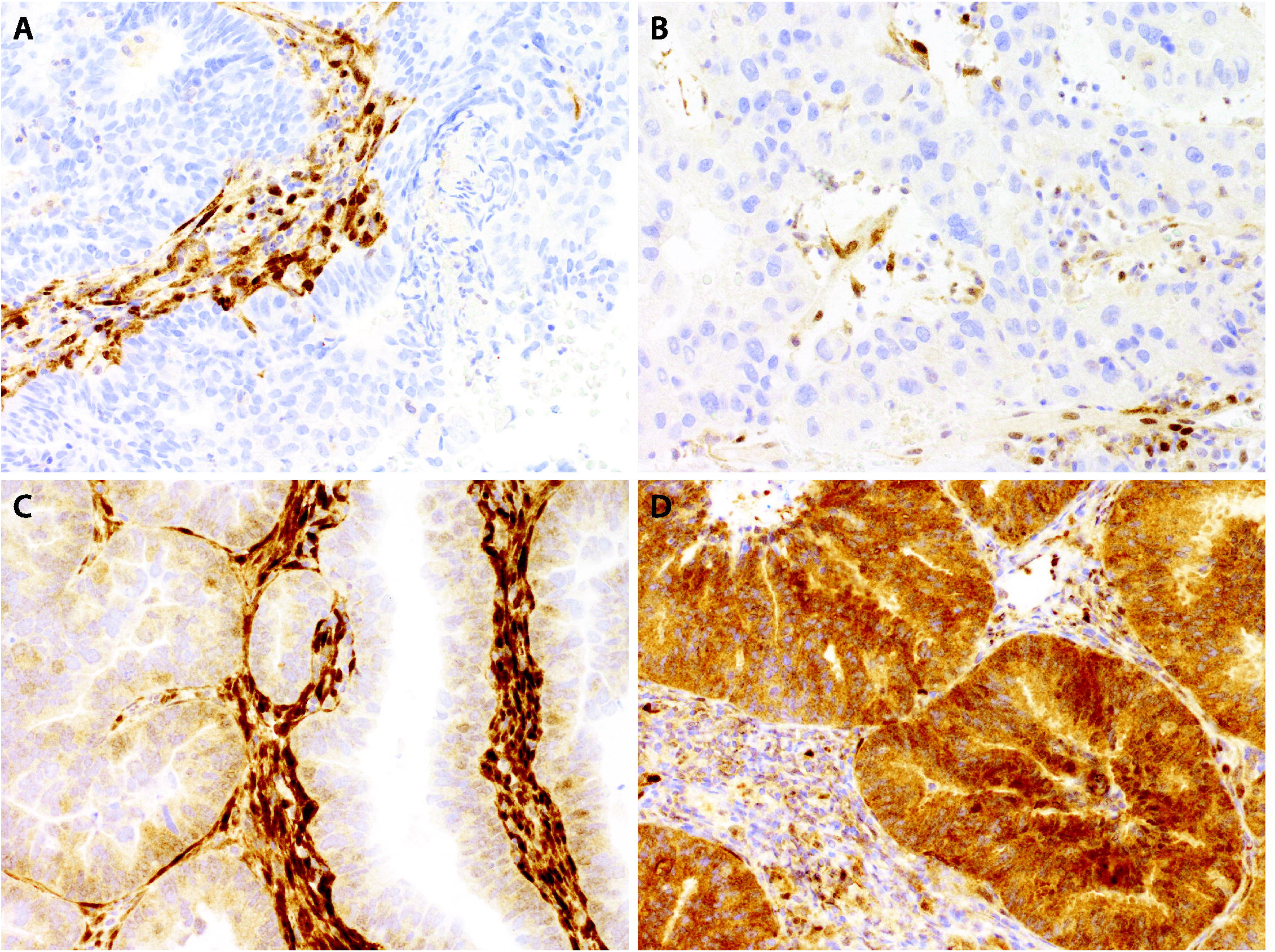
Immunohistochemistry staining pattern of PTEN. Absent (A), equivocal (B), reduced (C), and retained (D).

Because the sampling protocol for sequencing was not optimized to detect subclonal alterations, we excluded 17/182 (9.3%) cases with a subclonal IHC pattern from subsequent analyses (Figure 2). Still the agreement between *PTEN* mutation and IHC status was not perfect (Table 1). The sensitivity of absent PTEN IHC to predict a *PTEN* LOF mutation was 75.4% (95% CI 62.7 – 85.5%), the specificity was 84.6% (95% CI 76.2 – 90.9%), and the accuracy was 81.2% (74.4 – 86.9%). There were 16 cases where PTEN expression was absent but no LOF mutation was detected. The IHC patterns of discordant cases were re-reviewed but no revisions were made. On the other hand, there were 15 cases with retained PTEN expression in the presence of a LOF mutation. Detailed categorization on the LOF mutations of these 15 cases are shown in Table 2. 10 of 15 false positive cases could be explained by either low allelic frequency of the detected LOF mutation, late truncating mutation where mRNA is not undergoing nonsense mediated decay, or splicing mutation with unpredictable effect on expression and/or IHC failure. Notably, 11 out of 15 false positive cases harbored a second *PTEN* mutation (14/15 nonsynonymous, 1 splicing).

**Table 2:**
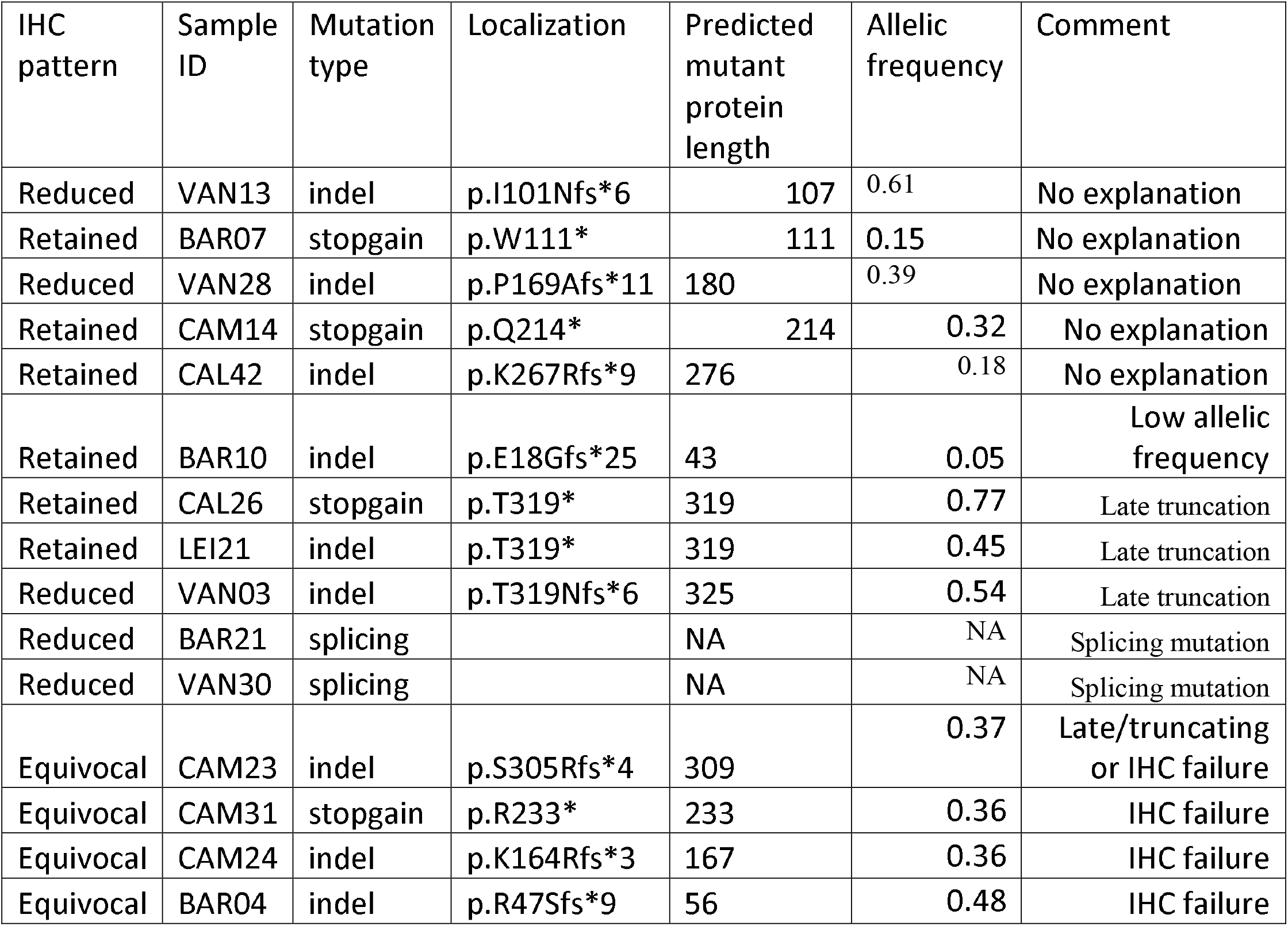
False negative IHC case with Loss-of-function (LOF) mutation and present PTEN expression by IHC.

**Figure 2.**
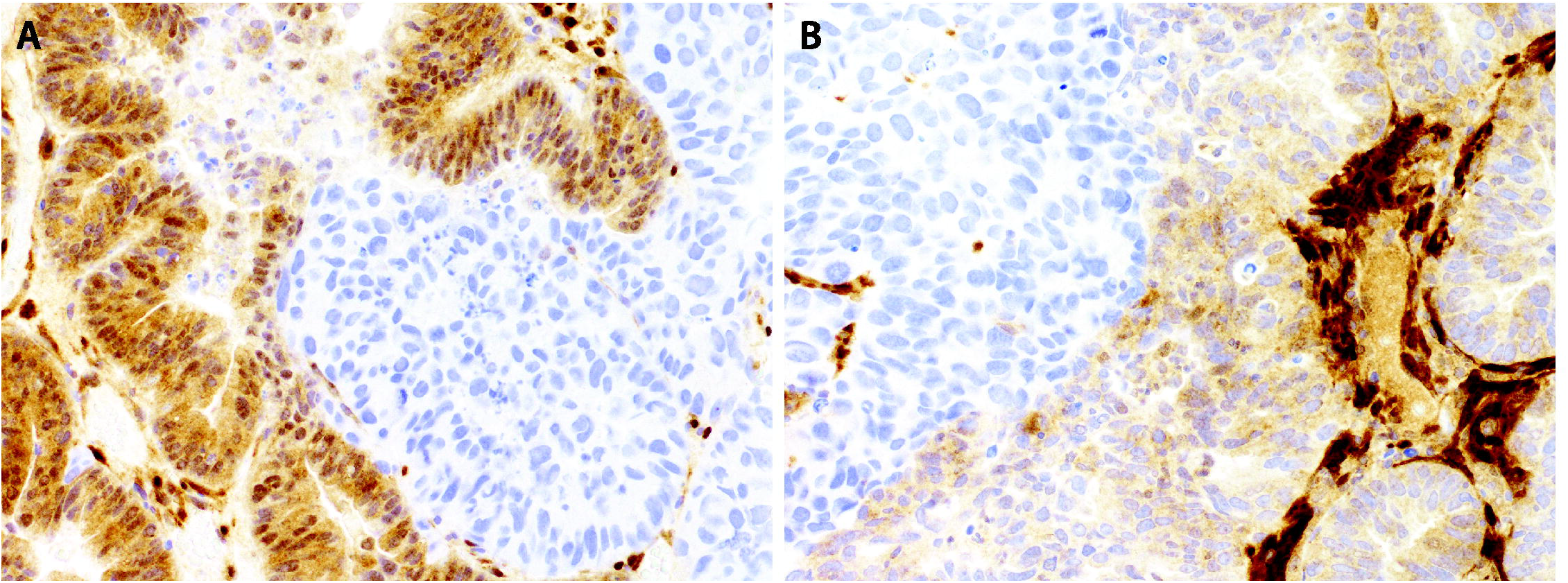
Subclonal staining pattern of PTEN. Case with retained staining and clearly demarcated absent area on the right (A), case with reduced staining in tumor cells compared to stronger intensity in stroma and absent staining on the left (B).

### *PTEN* Alterations by Histotype and Grade

We directly compared the result of the IHC staining pattern with sequencing analysis in determining PTEN abnormalities across histotype/grade categories (Table 3). Overall, sequencing detected more abnormalities than IHC (44.7% versus 37.6%). Because of the substantial number of discordant cases above we created the additional variable of abnormal PTEN by complementary interpretation of both assays, in which, abnormal PTEN includes absence of PTEN expression by IHC and/or presence of any LOF and NS *PTEN* mutation. Abnormal PTEN by this definition was present in 91.9% of endometrial endometrioid carcinoma, grade 1.The frequency of abnormal PTEN was significantly higher in endometrial endometrioid carcinomas of all grades compared to endometrial serous carcinoma (80.0% versus 19.4%, p<0.0001). Across centres, the frequency of abnormal PTEN ranged from 0 to 33% for endometrial serous and 70.8 to 100% for endometrial endometrioid carcinomas. Similarly, the frequency of *PTEN* mutations was significantly higher in endometrial endometrioid carcinomas of all grades compared to endometrial serous carcinoma (72.5% versus 13.6%, p<0.0001). Across centres, the frequency of *PTEN* mutations ranged from 0 to 28% for endometrial serous (2 centres with 0%) and 61.5% to 78.8% for endometrial endometrioid carcinomas.

**Table 3:** Comparison of PTEN alteration by IHC, sequencing and complementary interpretation across histotype/grade categories of endometrial carcinoma (cases with subclonal IHC or VUS by sequencing excluded).

|  | Absence of PTEN IHC | LOF or NS *PTEN* mutation | Abnormal PTEN by complementary interpretation |
| --- | --- | --- | --- |
| All cases, n (%) | 62/165 (37.6%) | 76/170 (44.7%) | 85/165 (51.5%) |
| EEC1, n (%) | 22/37 (59.5%) | 33/39 (84.6%) | 34/37(91.9%) |
| EEC2, n (%) | 10/22 (45.5%) | 14/18 (77.8%) | 16/22 (72.7%) |
| EEC3, n (%) | 13/21 (61.9%) | 11/23 (47.8%) | 14/21 (66.7%) |
| Undifferentiated, n (%) | 0/1 (0%) | 0/1 (0%) | 0/1 (0%) |
| Serous, n (%) | 9/62 (14.5%) | 9/66 (13.6%) | 12/62 (19.4%) |
| Mixed, n (%) | 5/9 (55.6%) | 7/10 (70%) | 6/9 (66.7%) |
| Carcinosarcoma, n (%) | 3/8 (37.5%) | 2/8 (25%) | 3/8 (37.5%) |
| Clear cell, n (%) | 0/5 (0%) | 0/5 (0%) | 0/5 (0%) |

### PTEN alterations by molecular subtype

The frequencies of absent PTEN staining, *PTEN* mutations and combined abnormal PTEN across the molecular subtypes of endometrial endometrioid carcinomas are shown in Table 4. Similar to previous published finding from the TCGA cohort, there is very high rate of abnormal PTEN in POLEmut and MMRd endometrial endometrioid carcinomas while the frequency within NSMP and p53abn subtypes are somewhat lower (non-significant trend, p=0.18). Subclonal PTEN loss was seen across all molecular subtypes with similar frequency.

**Table 4:** Comparison of PTEN alteration by IHC, sequencing and complementary interpretation across molecular subtypes of endometrial endometrioid carcinomas (cases with subclonal IHC or VUS by sequencing excluded).

|  | Absence of PTEN IHC | LOF or NS *PTEN* mutation | Abnormal PTEN by complementary interpretation |
| --- | --- | --- | --- |
| All EEC, n (%) | 45/80 (56.2%) | 58/80 (72.5%) | 64/80 (80%) |
| POLEmut, n (%) | 4/8 (50.0%) | 7/8 (87.5%) | 7/8 (87.5%) |
| NSMP, n (%) | 19/31 (61.3%) | 20/28 (71.4%) | 25/31 (80.7%) |
| MMRd, n (%) | 16/30 (53.3%) | 27/32 (84.4%) | 26/30 (86.7%) |
| P53abn, n (%) | 4/8 (50%) | 4/8 (50%) | 6/11 (54.5%) |

## DISCUSSION

Our study shows good agreement between PTEN IHC and LOF mutation in *PTEN*. However, the accuracy is not sufficiently high for PTEN IHC to be considered a surrogate for *PTEN* mutation. While some cases harboring a *PTEN* mutation still express PTEN by IHC (“false negative IHC”), there is a similar number of cases with unequivocal absence of PTEN protein expression that is likely caused by mechanisms other than mutations. We propose a combined assessment of both IHC and sequencing for a comprehensive interpretation of the PTEN status. The validity of this assessment is supported by the high frequency of PTEN alterations detected by this combined approach in endometrial endometrioid carcinomas.

Djordjevic and colleagues showed similar accuracy of PTEN IHC in predicting *PTEN* LOF mutations by Sanger sequencing in endometrial carcinoma: their accuracy was 73.2% (calculated from Table 6, excluding subclonal) compared to 81.2% in our study (9). Herein, we report a lower sensitivity (75.4% compared to 86.3% by Djordjevic et al.) but a higher specificity (84.6% compared to 64.5% by Djordjevic et al.) (9). The higher specificity is partly due to the higher sensitivity of NGS as compared to Sanger sequencing, i.e. a higher detection rate of mutations in IHC absent cases. Even though our cohort was selected in favor of endometrial carcinoma with *TP53* mutation, the overall frequency of *PTEN* mutation still holds to what has been reported previously (34- 65%) (13,17,18). We showed one of the highest frequencies of *PTEN* mutations in endometrial endometrioid carcinomas (72.5%), particularly grade 1 (84.6%), ever reported. Yet even with the superior sensitivity of NGS we still observed a relatively high number (N=16) of cases that showed unequivocal absence of PTEN expression but no detectable mutation. This is in line with previous observations that IHC detects a number of epigenetic abnormalities not seen by exome sequencing. Majority of the changes are attributable to *PTEN* promoter hypermethylation, which has been detected in 19% of endometrial carcinomas (19). In contrast, copy number loss (homozygous or heterozygous), which is present in 6% of tubo-ovarian high-grade serous carcinoma, does not seem to play a major role in endometrial carcinoma (only present in 2% of endometrial carcinomas in the TCGA dataset)(18). Furthermore, PTEN IHC can detect loss of PTEN expression by various other mechanisms, including microRNAs, regulation of protein stability and degradation (20).

The lower sensitivity of IHC to predict PTEN mutations in our study compared to Djordjevic and colleagues is due to 15 cases with detectable LOF mutation but unequivocally retained PTEN expression by IHC. Most of these cases are explained by the nature of the mutation detected: three cases with late truncating LOF mutations may produce a non-functional protein that can be still detected by IHC using an N-terminal antibody; three additional cases with splicing mutations may have an unpredictable effect on protein transcription; low allelic frequency was likely the cause in two cases. Only the remaining five cases may be related to the differences in the IHC assay between the two studies. Our study (clone 138G6 Cell signalling on DAKO Omnis) and the study by Djordevic et al. (Dako clone 6H2.1 on Leica Bond) used different IHC protocols (9). Both PTEN antibodies have been shown to correlate with PI3K-AKT activation (21-23). We noted some potential technical issues related to the assay during the interpretation, particularly with cases with low protein expression. Even after consolidating the results by discussing the interpretation at a multithreaded microscope, 6.6% of cases remained equivocal for PTEN loss (equivocal category). These cases had a cytoplasmic blush in the of tumor cells compared to the complete loss of staining in the “absent” category. Yet they are recognizably weaker compared to the “reduced/attenuated” category, which showed reduced staining in tumor cells compared to the on-slide internal control, but the tumor cells were unequivocally positive. When grouping equivocal with retained and subclonal loss with absent, the interobserver agreement in our study was slightly lower compared to a study using the Dako clone 6H2.1 (86% versus 91%).(8) This and the higher sensitivity to detect LOF mutations could be interpreted slightly in favor of the combination Dako clone 6H2.1 on a Leica Bond compared to Cell Signaling clone 138G6 on DAKO Omnis.(6)

Absent PTEN expression was throughout the entire tumor section in most of the cases, which is consistent with an early truncal event in the carcinogenesis of endometrial endometrioid carcinoma (2). However, a substantial proportion (9.3%) showed subclonal PTEN loss, indicative that some PTEN alterations occur as a later clonal event during tumor progression. The latter occurred in all molecular subtypes of endometrial endometrioid carcinoma and also in endometrial serous carcinomas. This pattern has been previously termed as heterogeneous (2,9). The loss of PTEN expression can be very focal, which stresses the importance of using whole paraffin sections for PTEN IHC. This subclonal/heterogeneous expression pattern of PTEN showed a significant discordance with the mutation status in our series. Targeted punches to areas with IHC loss would minimize this type of error for future sequencing analysis. A reduced pattern of PTEN expression has been previously described and has been historically grouped with complete loss of PTEN protein as abnormal (2). In our series, the majority of cases with reduced PTEN expression did not harbor LOF mutations (87.2%) similar to those with retained expression (88.5%). Thus, our data do not support grouping the reduced pattern with the absent pattern. The aforementioned issues plus the presence of 5% nonsynonymous *PTEN* mutations without change in PTEN expression, all lead to a lower total accuracy of PTEN IHC to predict LOF mutations compared to other proteins such as TP53 or ARID1A (24) (25). In both TP53 and ARID1A, a loss of protein function is rarely caused by epigenetic regulation.

Our findings have clinical relevance because PTEN has been proposed to aid in the diagnostic distinction of endometrioid versus serous histotype particularly in p53abn endometrial carcinomas (7,26). Notably, the frequency of *PTEN* mutations in endometrial serous carcinoma in our cohort is remarkably higher than in the TCGA dataset (13.6% versus 2.6%)(1). Given the issues with interobserver reproducibility in high-grade endometrial carcinoma (5,27), one explanation is difference in diagnostic standards across centres. This is corroborated by a wide range of *PTEN* mutation in endometrial serous carcinomas across centres: 2 centres had zero versus one center, which had a frequency of 28%. Further studies on the value of PTEN as a diagnostic aid in the distinction of p53abn endometrial endometrioid and serous carcinomas should be carefully designed. Another potential diagnostic application is the distinction of neoplastic from non-neoplastic proliferations in the endometrium. Increasing with older age, PTEN null glands by IHC can occur in histologically normal endometrium but these latent changes are not sufficient (not rate limiting) for neoplastic transformation (28-30). However, this concept has been recently challenged in that *PTEN* mutations were rarely found in histologically normal endometrium suggesting that PTEN may in fact represent a gatekeeper (31). However, the latter study did not use PTEN IHC for targeted laser microdissection of PTEN null glands as compared to the older studies (28,29). For practical purposes, absence of PTEN expression in isolated glands of histologically normal endometrium represents an unknown risk for progression and should therefore not be considered as a reportable finding (29). However, absence of PTEN expression in an architecturally crowded lesion with cytological demarcation can support the diagnosis of atypical endometrial hyperplasia as a precursor to endometrioid carcinoma.

In summary, we show that although IHC of PTEN expression provides a good initial assessment for PTEN status, neither IHC nor sequencing alone is sufficient to identify all cases with PTEN abnormality. We believe that complementary PTEN IHC and NGS should become standard for assessing the PTEN status particularly if this is for inclusion to clinical trials.

## Data Availability

on request

## Acknowledgement

We thank Shuhong Liu and Young Ou (Anatomical Pathology Research Laboratory, Department of Pathology) for immunohistochemical stains. MK received internal support from Calgary Laboratory Services (RS19-612). This project was funded by a research grant from Barts and the London Charity, MRD0206, supported by the Gynaecological Cancer Research Fund at Barts and the London NHS Trust.

